# Reassessing the link between adiposity and head and neck cancer: a Mendelian randomization study

**DOI:** 10.1101/2024.11.21.24317707

**Authors:** Fernanda Morales-Berstein, Jasmine Khouja, Mark Gormley, Elmira Ebrahimi, Shama Virani, James McKay, Paul Brennan, Tom G Richardson, Caroline L Relton, George Davey Smith, M Carolina Borges, Tom Dudding, Rebecca C Richmond

## Abstract

**Background:** Adiposity has been associated with an increased risk of head and neck cancer (HNC). Although body mass index (BMI) has been inversely associated with HNC risk among smokers, this is likely due to confounding. Previous Mendelian randomization (MR) studies could not fully discount causality between adiposity and HNC due to limited statistical power. Hence, we aimed to revisit this using the largest genome-wide association study (GWAS) of HNC available, which has more granular data on HNC subsites.

**Methods:** We assessed the genetically predicted effects of BMI (N=806,834), waist-to-hip ratio (WHR; N=697,734) and waist circumference (N=462,166) on the risk of HNC (N=12,264 cases and 19,259 controls) and its subsites (oral, laryngeal, hypopharyngeal and oropharyngeal cancers) using a two-sample MR framework. We used the inverse variance weighted (IVW) MR approach and multiple sensitivity analyses including the weighted median, weighted mode, MR-Egger, MR-PRESSO, and CAUSE approaches. We also used multivariable MR (MVMR) to explore the direct effects of the adiposity measures on HNC, while accounting for smoking behaviour, a well-known HNC risk factor.

**Results:** In univariable MR, higher genetically predicted BMI increased the risk of overall HNC (IVW OR=1.17 per 1 standard deviation [1-SD] higher BMI, 95% CI 1.02–1.34, p=0.03), with no heterogeneity across subsites (Q p=0.78). However, the effect was not consistent in sensitivity analyses. The IVW effect was attenuated when smoking was included in the MVMR model (OR accounting for comprehensive smoking index=0.96 per 1-SD higher BMI, 95% CI 0.80–1.15, p=0.64) and CAUSE indicated the IVW results could be biased by correlated pleiotropy. Furthermore, we did not find a link between genetically predicted WHR (IVW OR=1.05 per 1-SD higher WHR, 95% CI 0.89–1.24, p=0.53) or waist circumference and HNC risk (IVW OR=1.01 per 1-SD higher waist circumference, 95% CI 0.85–1.21, p=0.87).

**Conclusions:** Our findings suggest that adiposity does not play a major role in HNC risk.

## Introduction

Head and neck cancer (HNC) is among the ten most common cancers in Europe, with an age standardised incidence rate of 10.3 per 100,000 person-years[1]. Around 90% of HNCs are classed as squamous cell carcinomas of the oral cavity, pharynx or larynx[2]. Tobacco smoking and alcohol consumption are well-established HNC risk factors[3-9]. High-risk human papillomavirus (HPV) infection has also been causally linked to the risk of HNC, especially oropharyngeal cancer[10-12]. In contrast, the role of adiposity in the development of HNC is less well understood.

The World Cancer Research Fund/American Institute for Cancer Research (WCRF/AICR) Continuous Update Project (CUP) Expert Report published in 2018 determined higher body fatness (i.e., body mass index [BMI], waist-to-hip ratio [WHR] and waist circumference) likely increases the risk of HNC[13]. The CUP panel reached this conclusion even though a higher BMI has been associated with a decreased risk of HNC[13], since they noted the inverse association appears to be limited to current smokers. They concluded the association between BMI and HNC risk may be biased among individuals who smoke (because smoking is a HNC risk factor associated with lower weight). It is thought that nicotine consumption could lead to appetite suppression and increased energy expenditure, which could in turn, lead to weight loss (and spurious inverse associations between BMI and HNC risk[14]) among smokers[15]. Among never smokers, BMI has been positively associated with HNC risk, in line with the evidence observed for measures of central adiposity (i.e., WHR and waist circumference)[14, 16].

However, the relationship between adiposity and smoking is complex, with evidence from Mendelian randomization (MR) studies suggesting higher adiposity increases the risk of smoking[17, 18] while simultaneously suggesting smoking may lead to lower adiposity[17, 19-22]. Additionally, excess adiposity, socioeconomic deprivation and (both active and passive) smoking are often strongly correlated[23-26]. Thus, the positive associations between adiposity (i.e., BMI among non-smokers, WHR and waist circumference) and HNC risk may not be as unbiased as they appear.

It is important to acknowledge that previous MR studies on adiposity (i.e., BMI, WHR, waist circumference) and HNC risk were relatively small (maximum N=6,034 cases) and could not fully discount causality due to limited statistical power[27-29]. Therefore, the aim of this MR study was to revisit the link between adiposity and HNC risk using data from a HNC genome-wide association study (GWAS) that includes over two times the number of cases than the Genetic Associations and Mechanisms in Oncology (GAME-ON) GWAS[30] used by Gormley et al.[28] (the largest to date; N=12,619, including 6,034 cases and 6,585 controls) and has more granular data on HNC subsites (i.e., oral cavity, hypopharynx, oropharynx and larynx). We also aimed to use multivariable MR (MVMR) to explore the direct effects of the adiposity measures on HNC, while accounting for smoking behaviour.

## Methods

### Study design

We used a two-sample MR framework to assess the genetically predicted effects of BMI, WHR and waist circumference on the risk of HNC and its subsites (oral, laryngeal, hypopharyngeal and oropharyngeal cancers) among individuals of European ancestry. Genetic variants associated with these adiposity traits were used as instrumental variables to estimate causal effects under the three core MR assumptions[31]: 1) the genetic variants are strongly associated with the adiposity trait of interest (relevance assumption); 2) the distribution of the genetic variants in the population is not influenced by factors that also influence HNC risk, such as population stratification, assortative mating and dynastic effects (independence assumption); and 3) the genetic variants can only influence HNC risk via their effect on the adiposity trait of interest (exclusion restriction assumption). This work was conducted and reported according to the STROBE-MR guidelines[32] (**Additional File 1: Appendix**).

### Head and neck cancer GWAS

GWAS summary statistics for HNC were obtained from a European HEADSpAcE consortium GWAS that excluded UK Biobank participants (N=31,523, including 12,264 cases and 19,259 controls) to avoid overlapping samples across the exposure and outcome datasets. It includes the European GAME-ON data used by Gormley et al.[30] and has more granular data on HNC subsites (i.e., oral cavity [N=21,269, including 3,091 cases and 18,178 controls], hypopharynx [N=18,652, including 474 cases and 18,178 controls], HPV positive oropharynx [N=20,146, including 1,980 cases and 18,166 controls], HPV negative oropharynx [N=19,114, including 948 cases and 18,166 controls] and larynx [N=20,668, including 2,490 cases and 18,178 controls])[33].

HNC was defined based on the 10^th^ revision of the International Classification of Diseases (ICD-10)[34]. It included cancers of the oral cavity (C00.3, C00.4, C00.5, C00.6, C00.8, C00.9, C02.0–C02.9 [except C02.4], C03.0–C03.9, C04.0–C04.9, C05.0–C06.9 [except C05.1, C05.2]), the oropharynx (C01-C01.9, C02.4, C05.1, C05.2, and C09.0–C10.9), the hypopharynx (C12.0-C13.0), the larynx (C32), and overlapping or not otherwise specified sites (C14, C05.8, C02.8, C76.0)[33].

Further detail on the HEADSpAcE GWAS has been published elsewhere[33]. In brief, genotype data were obtained using nine different genotyping arrays. They were subsequently converted to genome build 38 for consistency across datasets. Quality control (QC) procedures were conducted by genotyping array rather than by study. Samples were excluded for the following reasons: sex mismatch (heterozygosity <0.8 for males and >0.2 for females), autosomal heterozygosity (>3 standard deviation [SD] units from the mean), missingness (>0.03), and cryptic relatedness (identity-by-decent >0.185). Single nucleotide polymorphisms (SNPs) were removed due to genotype missingness (>0.01), deviations from Hardy-Weinberg equilibrium (*p* <1e-05) and low minor allele count (<20). Imputation was performed using the TOPMed Imputation Server. Only SNPs with an imputation score r^2^ >0.3 and a minor allele frequency (MAF) >0.005 were included in the GWAS. The analyses were conducted in PLINK using logistic regressions adjusted for sex, the top principal components and imputation batch (six in total, which account for both genotyping array and study differences).

### Genetic instruments for adiposity

GWAS summary statistics for waist circumference (N=462,166) in SD units were obtained from the UK Biobank available via the IEU OpenGWAS platform (id: ukb-b-9405). GWAS summary statistics for BMI (N=806,834) and WHR (N=697,734) in SD units were obtained from the latest Genetic Investigation of Anthropometric Traits (GIANT) consortium’s GWAS meta-analysis by Pulit et al.[35] available at https://zenodo.org/records/1251813. The meta-analysis is the biggest to date, as it combines the meta-analysis by Shungin et al.[36] with UK Biobank data. The UK Biobank GWAS[35] was conducted using imputed data and the BOLT-LMM software[37]. The linear mixed models (LMMs) were solely adjusted for genotyping array. GIANT and UK Biobank data were meta-analysed[35] using an inverse-weighted fixed-effect meta-analysis in METAL[38].

We extracted GWAS-significant SNPs for waist circumference using the standard threshold (p<5e-08). For BMI and WHR, we extracted them according to the stringent threshold recommended by Pulit et al.[35] to account for denser imputation data (p<5e-09). We then performed LD-clumping to select independent lead SNPs for each exposure (r^2^=0.001, 10,000 kb). In total, 458 and 283 and 375 SNPs remained for BMI, WHR and waist circumference, respectively.

### Data harmonisation

We extracted HNC GWAS summary statistics that corresponded to the list of SNPs selected as instruments for the exposures. Proxy SNPs (r^2^>0.8) were used when the instrumental SNPs were not available in the outcome datasets. Proxies were identified using the “extract_outcome_data” function of the “TwoSampleMR” R package and the 1000 Genomes Project European reference panel. We harmonised the exposure and outcome datasets using the “harmonise_data” function of the “TwoSampleMR” R package[39]. Positive strands were inferred using allele frequencies and ambiguous palindromic SNPs with MAFs ≥0.3 were removed. The harmonised data used in the analyses are available in **Supplementary Tables 1 to 6 (Additional File 2)**.

We calculated mean F-statistics and total R^2^ values to assess the strength of our genetic instruments after data harmonisation[40, 41]. Consequently, we used the total R^2^ values to examine the statistical power in our study[42]. However, we acknowledge the value of post-hoc power calculations is limited, since the statistical power estimated for an observed association is already reflected in the 95% confidence interval presented alongside the point estimate[43].

### Statistical analysis

#### Main analyses

The multiplicative random effects inverse-variance weighted (IVW) MR approach[44] (the default IVW method of the “TwoSampleMR” package[39]) was used to investigate the genetically predicted effects of BMI, WHR and waist circumference on HNC risk. We did not correct our results for multiple testing, as all our exposures are strongly correlated[35, 36].

#### Sensitivity analyses

Because the IVW method assumes all genetic variants are valid instruments[44], which is unlikely the case, three pleiotropy-robust two-sample MR methods (i.e., MR-Egger[45], weighted median[46] and weighted mode[47]) were used in sensitivity analyses. When the magnitude and direction of effect estimates are consistent across methods that rely on different assumptions, the main findings are more convincing. As we cannot be sure about the presence and nature of horizontal pleiotropy, it is useful to compare results across methods even if they are not equally powered. We also performed tests for SNP heterogeneity (i.e., Q statistic test)[48] and directional horizontal pleiotropy (i.e., MR-Egger intercept test)[45]. When directional horizontal pleiotropy was identified, we used the intercept value to evaluate the extent of the bias. The MR-PRESSO[49] method was used to identify outliers (outlier test p<0.05) and calculate outlier-corrected causal estimates when there was evidence of SNP heterogeneity. The MR-PRESSO distortion test was used to evaluate differences between the outlier-corrected and IVW estimates.

In addition, we ran MVMR[50] analyses to evaluate the direct effects of adiposity measures with evidence of a total effect in our main analyses. The aim of the MVMR analyses was to separate the effect of adiposity from smoking behaviour (a well-known HNC risk factor which has a complex relationship with adiposity) in the development of HNC. We obtained genetic instruments for smoking behaviour from two different sources: a smoking initiation GWAS (N=805,431 excluding 23andme) derived by the GWAS and Sequencing Consortium of Alcohol and Nicotine use (GSCAN)[51] and a comprehensive smoking index (CSI; a measure of lifetime smoking that captures smoking heaviness, duration and cessation) GWAS (N=462,690) conducted by Wootton et al.[52]. Each was separately investigated in a MVMR framework. For each smoking trait, we selected SNPs that passed the GWAS-significance and independence thresholds (p<5e-08, r^2^=0.001, 10,000 kb) and combined them with the list of SNPs identified as instruments for the relevant exposure. We then performed LD-clumping across the combined list of SNPs, to then use these independent SNPs in MVMR analyses. The exposure and outcome datasets were harmonised to the same effect allele using the “harmonise_data” function of the “TwoSampleMR” R package[39]. We formatted the data using the “format_mvmr” function of the “MVMR” R package, calculated the conditional F-statistics for the MVMR instruments using the “strength_mvmr” function and ran the MVMR analyses using the “ivw_mvmr” function.

Since we used large GWAS datasets for the selection of our genetic instruments, our analyses are at an increased likelihood of being biased due to correlated horizontal pleiotropy (sometimes referred to as heritable confounding), which occurs when the genetic instruments are associated with the exposure through their effect on confounders of an exposure-outcome association[53, 54]. To mitigate this bias, we used Steiger filtering[55] to remove SNPs that are more strongly associated with smoking behaviour (a confounder of the exposure-outcome association) than the exposure of interest, as proposed by Sanderson et al.[56]. We also used Causal Analysis using Summary Effect Estimates (CAUSE)[54], another pleiotropy-robust MR method, to further investigate whether our results could be biased by correlated horizontal pleiotropy. CAUSE uses Bayesian expected log pointwise posterior predictive densities (ELPDs) to compare null, sharing and causal models. A higher ELPD represents a better model fit, so a positive delta ELPD (where delta ELPD = ELPD model 1 - ELPD model 2) suggests model 1 fits the data better than model 2, while a negative delta ELPD suggests the opposite. If we find evidence to reject the null hypothesis that the sharing model (i.e. causal effect fixed at zero) fits the data at least as well as the causal model (i.e. causal effect can differ from zero), our findings would be consistent with a causal effect. Steiger filtering and CAUSE analyses were only conducted for adiposity measures with evidence of a total effect in our main IVW analyses.

Moreover, we used the MR-Clust algorithm[57] to find distinct SNP clusters underlying the relationship between adiposity measures with evidence of a total effect in our main analyses and HNC. The identification of substantial clusters could provide insight into potential causal mechanisms. It could also flag pleiotropic variables that are associated with SNPs in each cluster. We filtered SNPs with conditional probabilities <0.8. At least four SNPs needed to remain per cluster for a substantial cluster to be reported.

#### Secondary analyses

In secondary analyses, we investigated the role of BMI, WHR and waist circumference on the risk of HNC by subsite (i.e., oral cavity, hypopharynx, HPV positive oropharynx, HPV negative oropharynx and larynx). We used a Cochran’s Q test to examine heterogeneity across HNC subsites.

We also explored the role of other adiposity-related anthropometric measures on the risk of HNC and its subsites. These anthropometric measures included: 1) four body shape principal components[58], 2) childhood and adulthood body size[59], 3) metabolically favourable and unfavourable adiposity[60], 4) body fat percentage, and 5) brain and adipose tissue-specific BMI[61]. The data sources for these traits are summarised in **Table 1**.

**Table 1.**
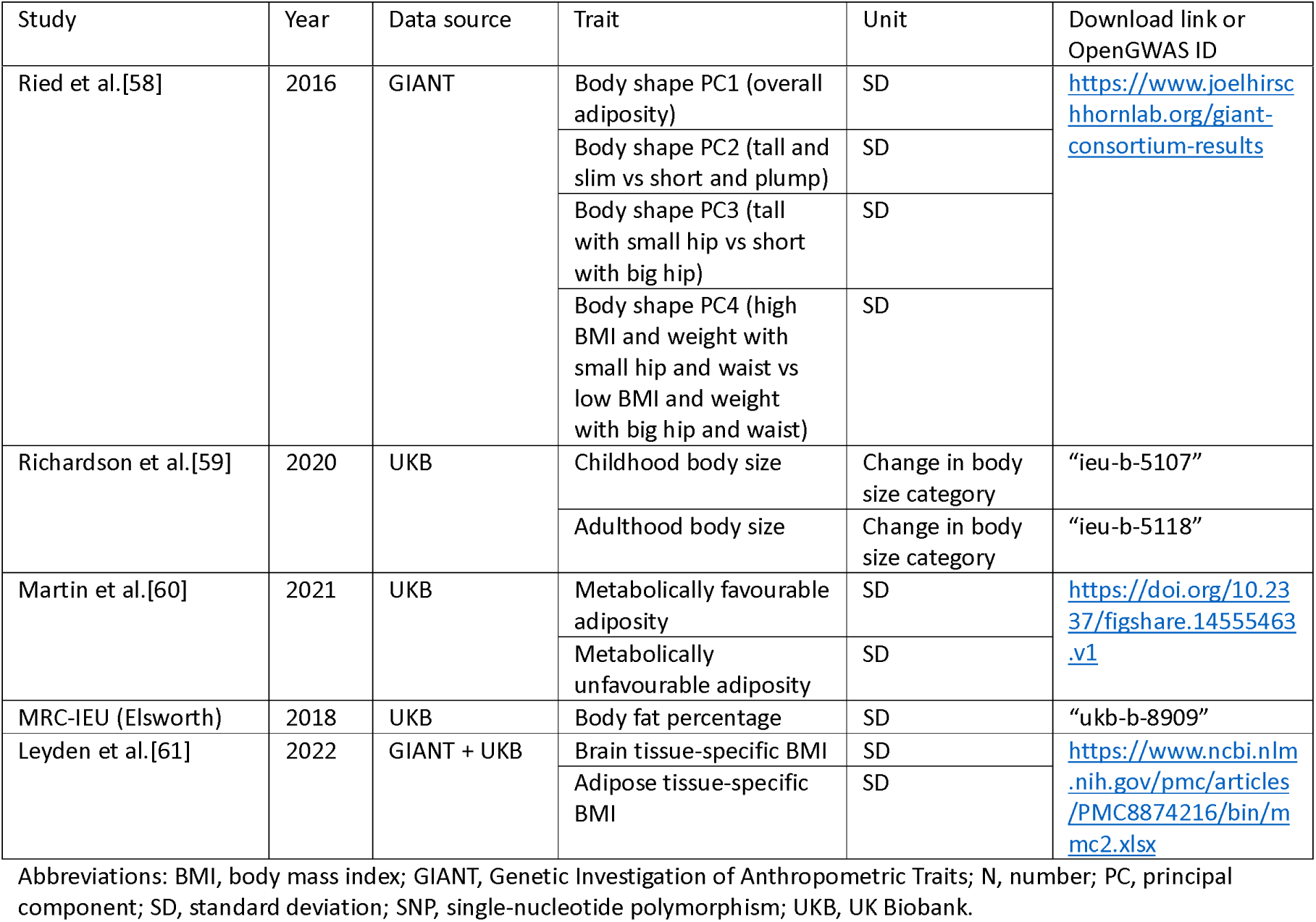
Data sources and instruments for other adiposity-related anthropometric measures.

#### Statistical software

We completed all MR analyses using R software version 4.4.0 and the “TwoSampleMR” v0.6.3, “MRPRESSO” v1.0, “MVMR” v0.4, “cause” v1.2.0 and “mrclust” v0.1.0 R packages. The “ggplot2” v3.5.1 and “ggforestplot” v0.1.0 R packages were used to create forest plots. The code used to run the MR analyses is available at http://github.com/fernandam93/adiposity_HNC_MR.

## Results

### Genetic instruments for BMI, WHR and waist circumference

After data harmonisation and the removal of ambiguous palindromic SNPs, 442 genetic variants remained as instruments for BMI, while 267 remained for WHR and 353 for waist circumference (**Additional File 2: Supplementary Table 1**). The mean F-statistic for BMI was 77 (range 33–844) and the total variance explained was 4.8%. For WHR, the mean F-statistic was 73 (range 33–820) and the total variance explained was 3.1%. For waist circumference, the mean F-statistic was 58 (range 30–940) and the total variance explained was 4.4%.

Using the BMI genetic instruments (total R^2^= 4.8%) and an α of 0.05, we had 80% statistical power to detect an OR as small as 1.16 for HNC risk (**Supplementary** Figure 1). For WHR (total R^2^= 3.1%) and WC (total R^2^= 4.4%), we could detect odds ratios (ORs) as small as 1.20 and 1.17, respectively. This is an improvement in terms of statistical power compared to the GAME-ON analysis published by Gormley et al.[28], for which there was 80% power to detect an OR as small as 1.26 using the same BMI genetic instruments (**Supplementary** Figure 2).

The F-statistics and R^2^ values for the other adiposity-related anthropometric measures have been summarised in **Table 2**.

**Table 2.**
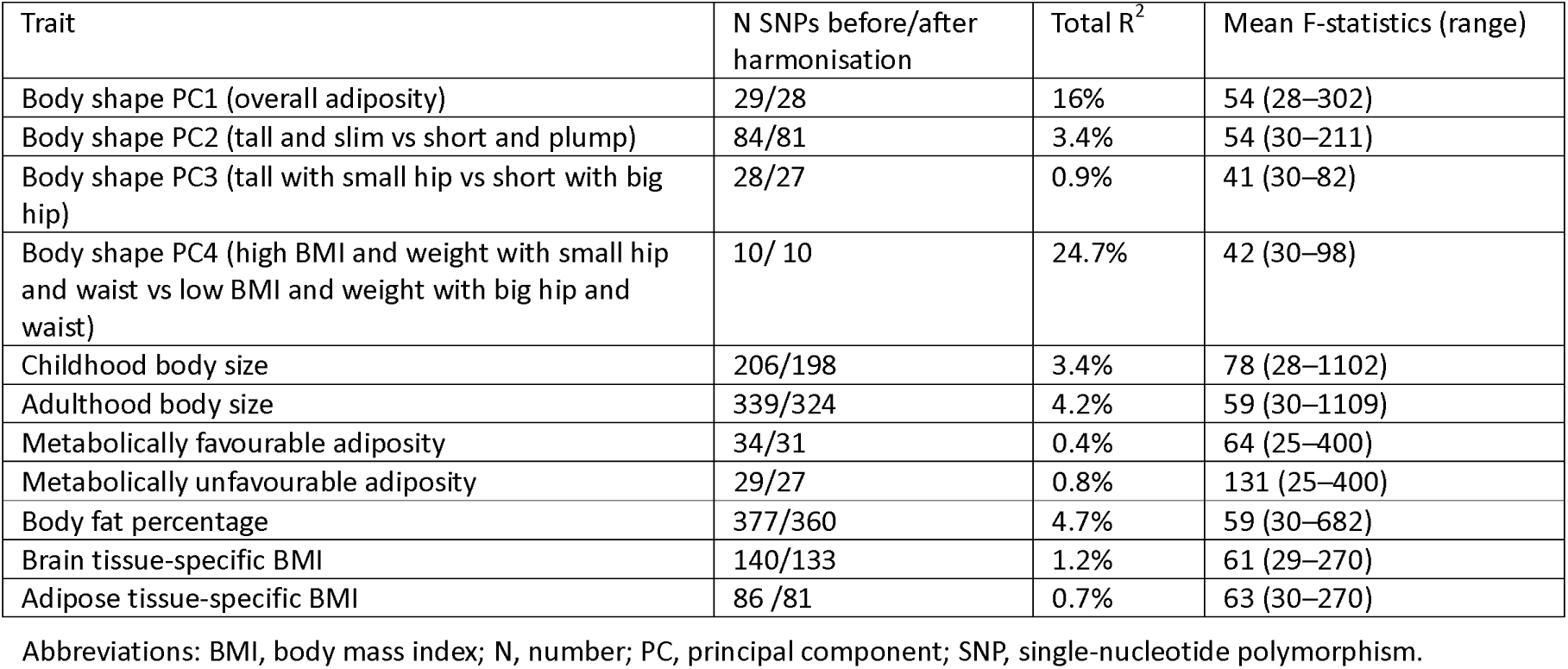
F-statistics and variance explained for other adiposity-related anthropometric measures.

| Trait | N SNPs before/after harmonisation | Total $R^2$ | Mean F-statistics (range) |
| --- | --- | --- | --- |
| Body shape PC1 (overall adiposity) | 29/28 | 16% | 54 (28–302) |
| Body shape PC2 (tall and slim vs short and plump) | 84/81 | 3.4% | 54 (30–211) |
| Body shape PC3 (tall with small hip vs short with big hip) | 28/27 | 0.9% | 41 (30–82) |
| Body shape PC4 (high BMI and weight with small hip and waist vs low BMI and weight with big hip and waist) | 10/ 10 | 24.7% | 42 (30–98) |
| Childhood body size | 206/198 | 3.4% | 78 (28–1102) |
| Adulthood body size | 339/324 | 4.2% | 59 (30–1109) |
| Metabolically favourable adiposity | 34/31 | 0.4% | 64 (25–400) |
| Metabolically unfavourable adiposity | 29/27 | 0.8% | 131 (25–400) |
| Body fat percentage | 377/360 | 4.7% | 59 (30–682) |
| Brain tissue-specific BMI | 140/133 | 1.2% | 61 (29–270) |
| Adipose tissue-specific BMI | 86 /81 | 0.7% | 63 (30–270) |

Abbreviations: BMI, body mass index; N, number; PC, principal component; SNP, single-nucleotide polymorphism.

### Genetically predicted effects of BMI, WHR and waist circumference on HNC risk

In univariable MR, higher genetically predicted BMI increased the risk of overall HNC (IVW OR=1.17 per 1 standard deviation [1-SD] higher BMI, 95% CI 1.02–1.34, p=0.03), with no heterogeneity across subsites (Q p=0.78) (**Figure 1, Supplementary** Figure 3 **and Additional File 2: Supplementary Table 7**). However, the positive relationship between genetically predicted BMI and HNC risk was not consistent across the MR-Egger, weighted median and weighted mode analyses, with point estimates in opposing directions and confidence intervals including the null. The Q statistic and MR-Egger intercept tests suggested that there was heterogeneity across individual SNP estimates (Q=609, p<0.001) and a minor degree of unbalanced horizontal pleiotropy (intercept=0.007, p=0.03) that could have biased the main IVW results (**Additional File 2: Supplementary Tables 8 and 9**). Although the MR-PRESSO analysis identified two outliers (i.e., rs11611246 and rs9603697), the distortion test suggested the outlier-corrected estimate (outlier-corrected IVW OR=1.14 per 1-SD higher BMI, 95% CI 1.00–1.30, p=0.05) was not statistically different to the main IVW estimate (p=0.94) (**Additional File 2: Supplementary Table 10**).

**Figure 1.**
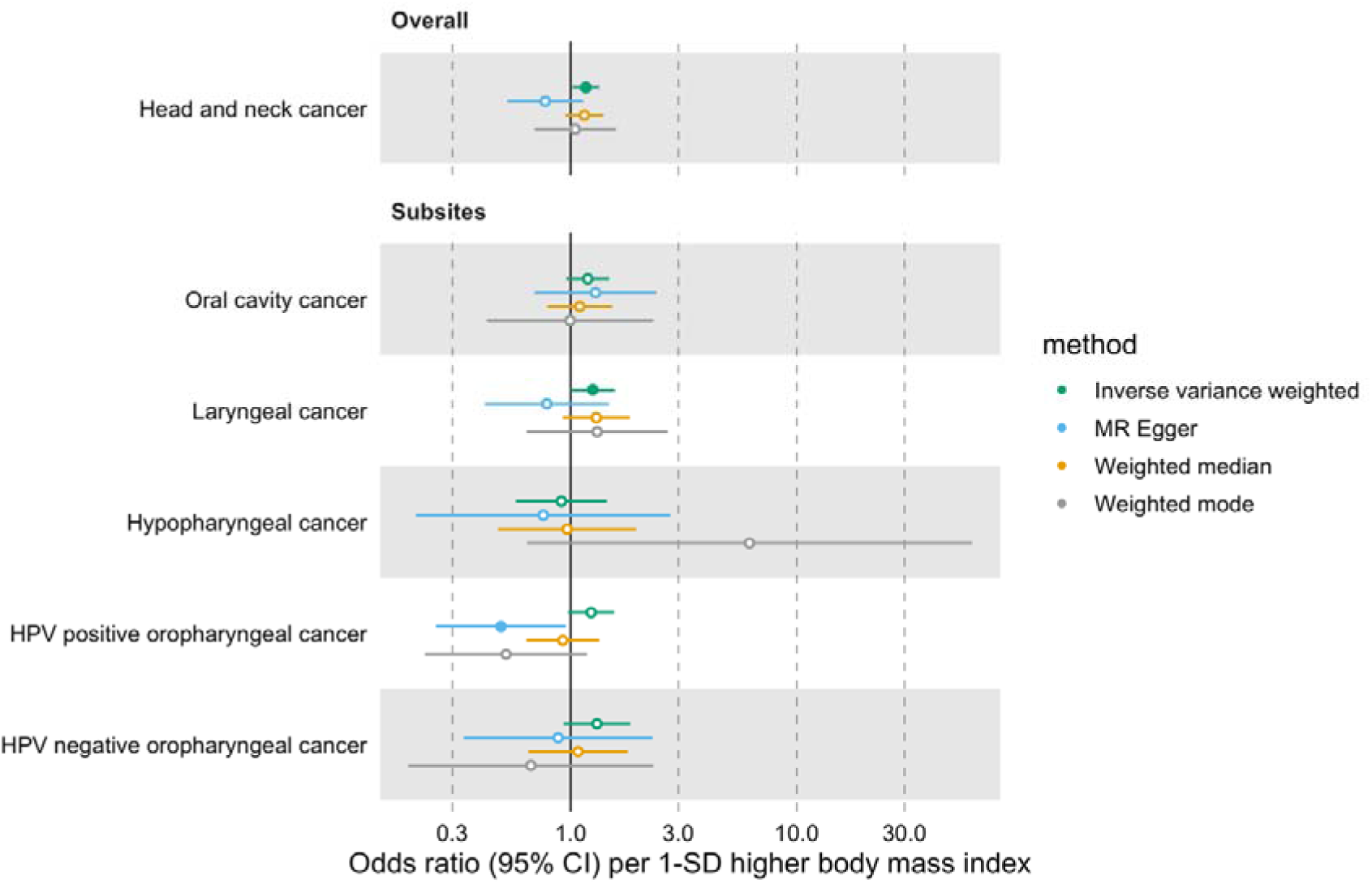
Forest plot for the genetically predicted effects of body mass index on the risk of head and neck cancer and its subsites.

Furthermore, we did not find a link between genetically predicted WHR and HNC risk (IVW OR=1.05 per 1-SD higher WHR, 95% CI 0.89–1.24, p=0.53) and there was no heterogeneity across subsites (Q p=0.15) (**Figure 2, Supplementary** Figure 4 **and Additional File 2: Supplementary Table 7**). MR-Egger, weighted median and weighted mode results were consistent with a null effect. The Q statistic and MR-Egger intercept tests suggested that there was some evidence of SNP heterogeneity (Q=332, p=0.004) and unbalanced horizontal pleiotropy (intercept=0.008, p=0.03) (**Additional File 2: Supplementary Tables 8 and 9**). The MR-PRESSO analysis did not identify any significant outliers (**Additional File 2: Supplementary Table 10**).

**Figure 2.**
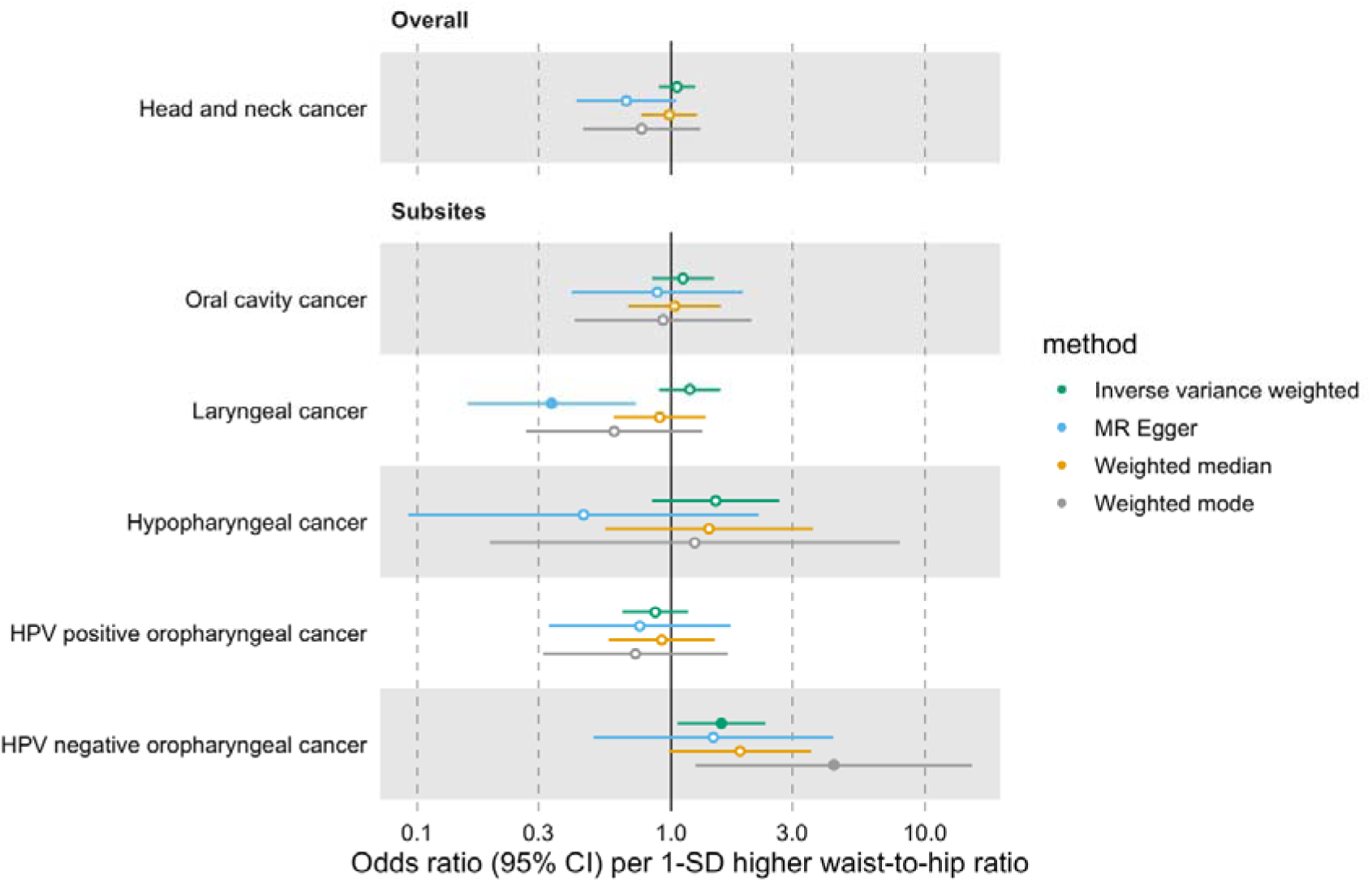
Forest plot for the genetically predicted effects of waist-to-hip ratio on the risk of head and neck cancer and its subsites.

Similarly, we did not find a genetically predicted effect of waist circumference on HNC risk (IVW OR=1.01 per 1-SD higher waist circumference, 95% CI 0.85–1.21, p=0.87) or evidence of heterogeneity across subsites (Q p=0.87) (**Figure 3, Supplementary** Figure 5 **and Additional File 2: Supplementary Table 7**). The MR-Egger, weighted median and weighted mode consistently suggested the absence of an effect of waist circumference on HNC risk. The Q statistic and MR-Egger intercept tests suggested SNP heterogeneity (Q=563, p<0.001) but no unbalanced horizontal pleiotropy (intercept=-0.002, p=0.68) (**Additional File 2: Supplementary Tables 8 and 9**). The MR-PRESSO analysis identified four outliers (i.e., rs1229984, rs1336486, rs17446091, and rs55726687) but the distortion test suggested the outlier-corrected estimate (outlier-corrected IVW OR=0.98 per 1-SD higher waist circumference, 95% CI 0.84–1.15, p=0.82) was not statistically different to the main IVW estimate (p=0.12) (**Additional File 2: Supplementary Table 10**).

**Figure 3.**
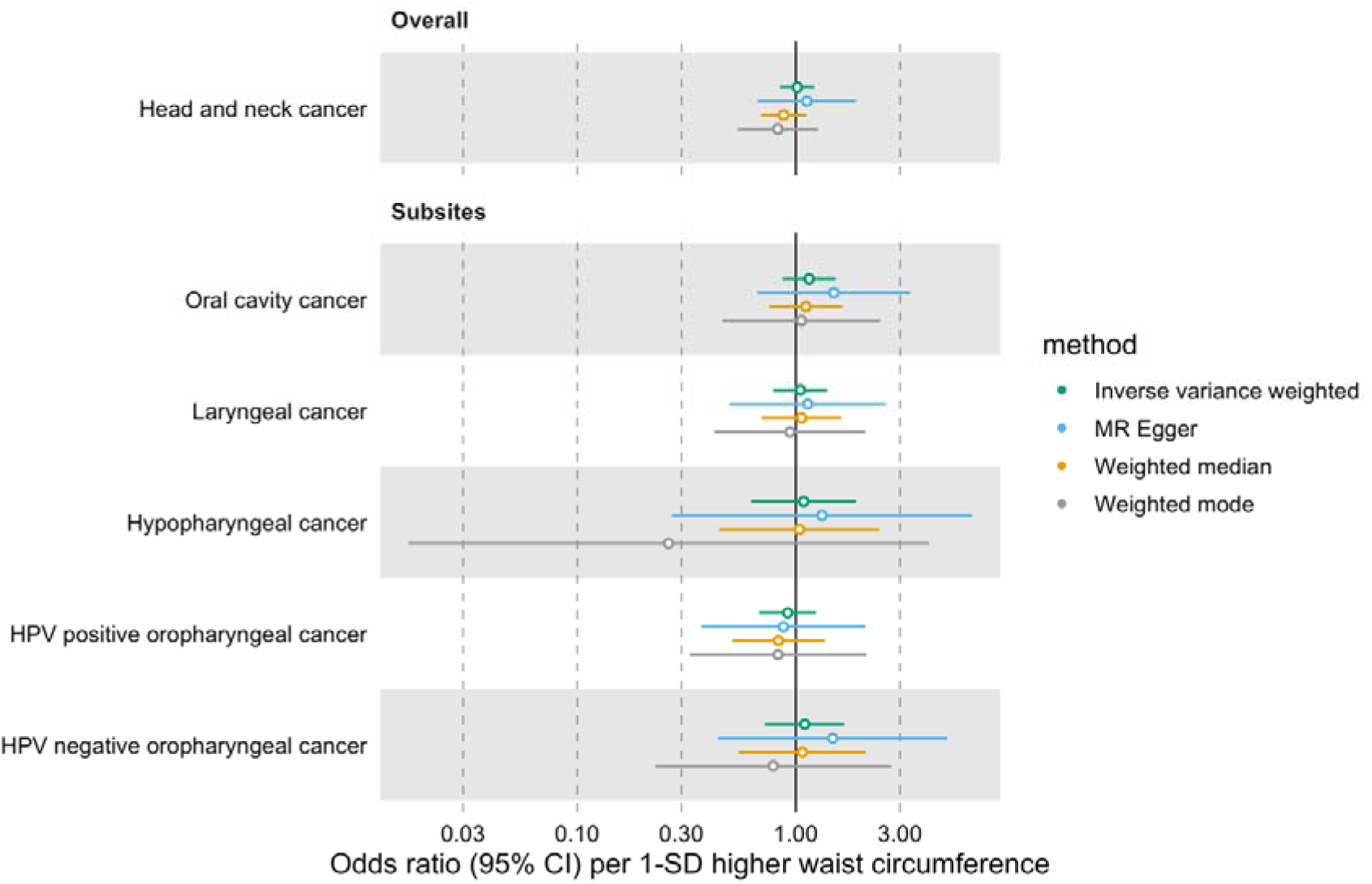
Forest plot for the genetically predicted effects of waist circumference on the risk of head and neck cancer and its subsites.

### MVMR estimates for BMI on HNC risk after accounting for smoking behaviour

In univariable IVW MR, both CSI and SI were linked to an increased risk of HNC (CSI OR=4.47 per 1-SD higher CSI, 95%CI 3.31–6.03, p<0.001; SI OR=2.07 per 1-SD higher SI 95%CI 1.60–2.68, p<0.001) (**Additional File 2: note in Supplementary Table 11**).

The effect of BMI on HNC risk was attenuated when smoking behaviour was included in the MVMR model (OR accounting for CSI=0.93 per 1-SD higher BMI, 95% CI 0.78–1.12, p=0.47; OR accounting for smoking initiation=1.09 per 1-SD higher BMI, 95% CI 0.88–1.34, p=0.43) (**Figures 4 and 5 and Additional File 2: Supplementary Table 11**). Genetically predicted smoking behaviour increased the risk of HNC even after accounting for BMI (CSI OR accounting for BMI=4.25 per 1-SD higher CSI, 95% CI 3.18–5.67, p<0.001; SI OR accounting for BMI=2.10 per 1-SD higher SI, 95% CI 1.61–2.73, p<0.001). The conditional F-statistics for the BMI estimates were 30.5 and 30.3 in the CSI and smoking initiation analyses, respectively (**Additional File 2: Supplementary Table 11**). They were slightly lower for the smoking behaviour estimates conditioning on BMI (13.4 and 19.5 in the CSI and smoking initiation analyses, respectively).

**Figure 4.**
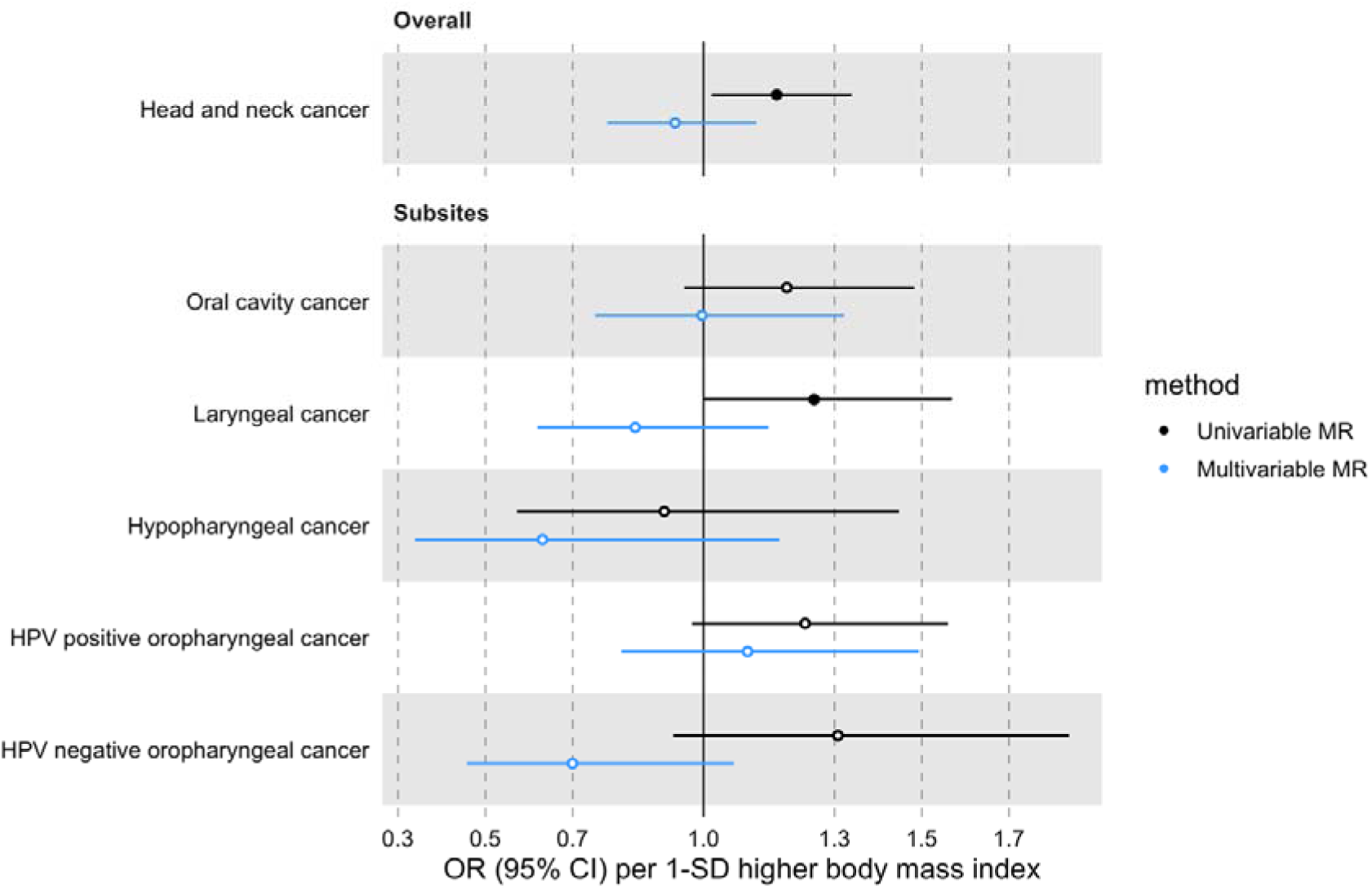
Forest plot for the genetically predicted effects of BMI on the risk of HNC and its subsites, before (univariable-black) and after (multivariable-blue) accounting for comprehensive smoking index (CSI).

**Figure 5.**
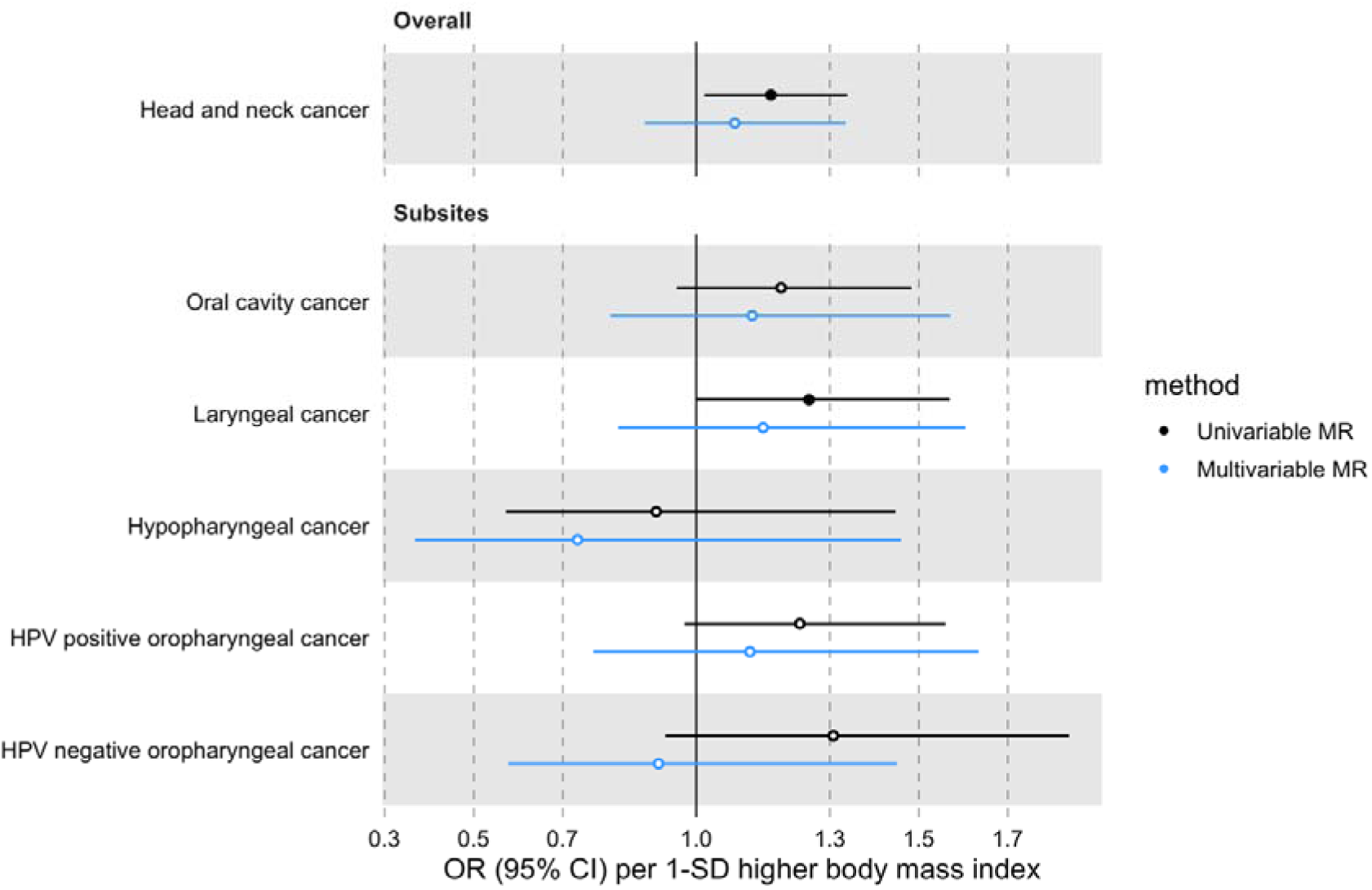
Forest plot for the genetically predicted effects of BMI on the risk of HNC and its subsites, before (univariable-black) and after (multivariable-blue) accounting for smoking initiation (SI).

### MR estimate for BMI on HNC risk after Steiger filtering SNPs more strongly associated with smoking behaviour than BMI

After removing six SNPs (i.e., rs10002111, rs2503185, rs264941, rs10858334, rs225882, rs2273175) that were more strongly associated with smoking behaviour (i.e., CSI or smoking initiation) than BMI, the genetically predicted effect of BMI on HNC risk slightly attenuated towards the null (Steiger filtered IVW OR=1.14 per 1-SD higher BMI, 95% CI 1.00–1.31, p=0.05) (**Additional File 2: Supplementary Table 12**).

### CAUSE estimate for BMI on HNC risk

We did not find evidence against bias due to correlated pleiotropy, since the causal model did not fit the data much better than the sharing model (CAUSE OR 1.12 per 1-SD higher BMI, 95% credible interval 0.93–1.34, delta ELPD for sharing vs causal=-0.07, *p*=0.47) (**Supplementary** Figure 6). Interestingly, neither the sharing nor the causal model fitted the data much better than the null model (delta ELPD for null vs sharing=-0.39, *p*=0.36; and delta ELPD for null vs causal=-0.46, *p*=0.41).

### MR-Clust estimates for the relationship between BMI and HNC risk

After filtering SNPs with conditional probabilities <0.8 and clusters with fewer than four SNPs (e.g., cluster 1, as only three of 17 SNPs remained after probability filtering), only a null cluster including 372 SNPs (424 before filtering) remained in the MR-Clust output for BMI and HNC risk (**Supplementary** Figure 7 **and Additional File 2: Supplementary Table 13**). Hence, the MR-Clust analysis did not reveal any mechanistic pathways underlying the effect observed.

### Genetically predicted effects of other adiposity-related anthropometric measures on HNC risk

We did not find consistent evidence of genetically predicted effects of other anthropometric measures on HNC risk (**Supplementary** Figures 8–12 **and Additional File 2: Supplementary Table 14**). The IVW estimate for PC2 capturing a combination of taller height and slimmer waist suggested this body shape decreased HNC risk (OR=0.86, 95% CI 0.75–0.99, p=0.04) (**Supplementary** Figure 8b). Similarly, the IVW estimate for PC3 capturing a combination of taller height and narrower hips suggested this body shape also reduced HNC risk (OR=0.73, 95% CI 0.55–0.97, p=0.03) (**Supplementary** Figure 8c). However, these inverse relationships were not consistent with results obtained using pleiotropy-robust methods (i.e., MR-Egger, weighted median and weighted mode).

## Discussion

In this MR study, we reaffirmed that there is no clear evidence of a genetically predicted effect of adiposity (i.e., BMI, WHR and waist circumference) or related anthropometric measures on the risk of HNC or its subsites. Although we found higher genetically predicted BMI increased the risk of overall HNC in the main univariable MR analysis, this was not consistent across the sensitivity analyses. Notably, the MVMR results suggested the main analysis may have been biased by smoking (and/or related traits), as the effect disappeared after accounting for smoking behaviour. The results obtained after Steiger filtering SNPs more strongly associated with smoking behaviour than BMI suggested correlated pleiotropy may have been partly biasing the BMI-HNC estimate. CAUSE, which is more robust to correlated pleiotropy than the IVW method, further supported this hypothesis.

Previous MR studies suggest adiposity does not influence HNC risk[27-29]. Gormley et al.[28] did not find a genetically predicted effect of adiposity on combined oral and oropharyngeal cancer when investigating either BMI (OR=0.89 per 1-SD, 95% CI 0.72–1.09, p=0.26), WHR (OR=0.98 per 1-SD, 95% CI 0.74–1.29, p=0.88) or waist circumference (OR=0.73 per 1-SD, 95% CI 0.52–1.02, p=0.07) as risk factors. Similarly, a large two-sample MR study by Vithayathil et al.[29] including 367,561 UK Biobank participants (of which 1,983 were HNC cases) found no link between BMI and HNC risk (OR=0.98 per 1-SD higher BMI, 95% CI 0.93–1.02, p=0.35). Larsson et al.[27] meta-analysed Vithayathil et al.’s[29] findings with results obtained using FinnGen data to increase the sample size even further (N=586,353, including 2,109 cases), but still did not find a genetically predicted effect of BMI on HNC risk (OR=0.96 per 1-SD higher BMI, 95% CI 0.77–1.19, p=0.69). With a much larger sample (N=31,523, including 12,264 cases), our IVW MR analysis suggested BMI may play a role in HNC risk, in contrast to previous studies. However, our sensitivity analyses implied that causality was uncertain.

In our study, we found some evidence that the genetically predicted effect of BMI on HNC risk was influenced by smoking. This could be due to the bidirectional relationship between smoking and adiposity reported in previous MR studies[17-22] or due to their shared genetic architecture[62, 63]. A strength of our study was that it was the first to exploit MVMR to disentangle the effects of BMI and smoking behaviour on the risk of HNC and its subsites. An advantage of our approach compared to conducting univariable MR analyses stratified by smoking status is that the former does not induce collider bias and provides estimates of direct effects irrespective of horizontal pleiotropy or mediation. Yet, we acknowledge the smoking behaviour traits used in our MVMR analyses likely capture more than just smoking, since some of the SNPs used to instrument these traits have been associated with risk-taking phenotypes and socioeconomic factors[64-67]. This places limits in the inferences that can be made about smoking in the context of mediation.

An important strength of our study was that the HEADSpAcE consortium GWAS used had a large sample size which conferred more statistical power to detect effects of adiposity on HNC risk compared to previous MR analyses[27-29]. Furthermore, the availability of data on more HNC subsites, including oropharyngeal cancers by HPV status, allowed us to investigate the relationship between adiposity and HNC risk in more detail than previous MR studies which limited their subsite analyses to oral cavity and overall oropharyngeal cancers[28, 68]. This is relevant because distinct HNC subsites are known to have different aetiologies[69], although we did not find evidence of heterogeneity across subsites in our analyses investigating the genetically predicted effects of BMI, WHR and WC on HNC risk.

We acknowledge that a major limitation of MR studies, including ours, is that several untestable assumptions are required to make accurate causal inferences. It is unlikely that our findings were influenced by weak instrument bias (i.e., violating the relevance assumption) because we used strong genetic instruments to proxy our adiposity traits. However, the independence assumption of no genetic confounding and the exclusion restriction assumption of no horizontal pleiotropy could have been violated. Furthermore, we were unable to explore potential non-linear causal effects of adiposity on HNC risk in the present study.

While our study contributes valuable evidence on the role of adiposity in the development of HNC, we recognise there is a need for additional research on the subject. Our study was limited to individuals of European ancestry, so our findings should be replicated in other ancestry groups before being generalised to non-European populations. Moreover, further research is needed to understand the biology underlying the complex relationship between smoking and adiposity, especially since it may be difficult to intervene on one without influencing the other[17].

## Conclusions

In conclusion, this study indicates that adiposity does not play a major role in HNC risk. Although we did not find strong evidence of a causal effect of adiposity on HNC, obesity is an established risk factor for multiple cancers and other chronic diseases[27, 70, 71]. Hence there is still value in aiming to reduce the levels of excess adiposity in the population.

## Data Availability

All the GWAS datasets used in our study are publicly available. The GWAS summary statistics for waist circumference are available via the IEU OpenGWAS platform (id: ukb-b-9405). The GWAS summary statistics for BMI and WHR by Pulit et al.[35] can be downloaded from https://zenodo.org/records/1251813. The data sources for the other adiposity-related measures have been specified in Table 1. The smoking behaviour traits GWAS data were downloaded from https://data.bris.ac.uk/data/dataset/10i96zb8gm0j81yz0q6ztei23d (for CSI) and https://doi.org/10.13020/przg-dp88 (for smoking initiation). The outcome datasets used in our analyses have been uploaded to the IEU OpenGWAS project platform for reproducibility. However, because the data was originally in build GRCh38, some multiallelic SNPs that could not be aligned with GRCh37 Human Genome reference sequence were dropped when lifting the data to build HG19/GRCh37 (which was required at the time of upload: April 2024). The following IEU OpenGWAS ids were assigned to the European HEADSpAcE HNC GWAS datasets including/excluding UK Biobank: ieu-b-5129/ieu-b-5123 for overall HNC, ieu-b-5132/ieu-b-5126 for oral cavity cancer, ieu-b-5130/ieu-b-5124 for hypopharynx cancer, ieu-b-5134/ieu-b-5128 for HPV positive oropharynx cancer, ieu-b-5133/ieu-b-5127 for HPV negative oropharynx cancer, and ieu-b-5131/ieu-b-5125 for larynx cancer. The R code used to run the MR analyses is available at http://github.com/fernandam93/adiposity_HNC_MR.

https://data.bris.ac.uk/data/dataset/10i96zb8gm0j81yz0q6ztei23d

https://gwas.mrcieu.ac.uk

## Availability of data and materials

All the GWAS datasets used in our study are publicly available. The GWAS summary statistics for waist circumference are available via the IEU OpenGWAS platform (id: ukb-b-9405). The GWAS summary statistics for BMI and WHR by Pulit et al.[35] can be downloaded from https://zenodo.org/records/1251813. The data sources for the other adiposity-related measures have been specified in Table 1. The smoking behaviour traits GWAS data were downloaded from https://data.bris.ac.uk/data/dataset/10i96zb8gm0j81yz0q6ztei23d (for CSI) and https://doi.org/10.13020/przg-dp88 (for smoking initiation). The outcome datasets used in our analyses have been uploaded to the IEU OpenGWAS project platform for reproducibility. However, because the data was originally in build GRCh38, some multiallelic SNPs that could not be aligned with GRCh37 Human Genome reference sequence were dropped when lifting the data to build HG19/GRCh37 (which was required at the time of upload: April 2024). The following IEU OpenGWAS id’s were assigned to the European HEADSpAcE HNC GWAS datasets including/excluding UK Biobank: ieu-b-5129/ieu-b-5123 for overall HNC, ieu-b-5132/ieu-b-5126 for oral cavity cancer, ieu-b-5130/ieu-b-5124 for hypopharynx cancer, ieu-b-5134/ieu-b-5128 for HPV positive oropharynx cancer, ieu-b-5133/ieu-b-5127 for HPV negative oropharynx cancer, and ieu-b-5131/ieu-b-5125 for larynx cancer. The R code used to run the MR analyses is available at http://github.com/fernandam93/adiposity_HNC_MR.

## Acknowledgements

We thank Richard Wilkinson for proofreading several versions of the manuscript. We would also like to thank Weili (Jason) Qiu, the IEU Data Manager, for his help debugging code and uploading the HNC GWAS summary statistics to the IEU OpenGWAS platform.

## IARC disclaimer

Where authors are identified as personnel of the International Agency for Research on Cancer/World Health Organization, the authors alone are responsible for the views expressed in this article and they do not necessarily represent the decisions, policy or views of the International Agency for Research on Cancer/World Health Organization

