## Additional file 3 for "Reassessing the link between adiposity and head and neck cancer: a Mendelian randomization study"

### Supplementary Figures

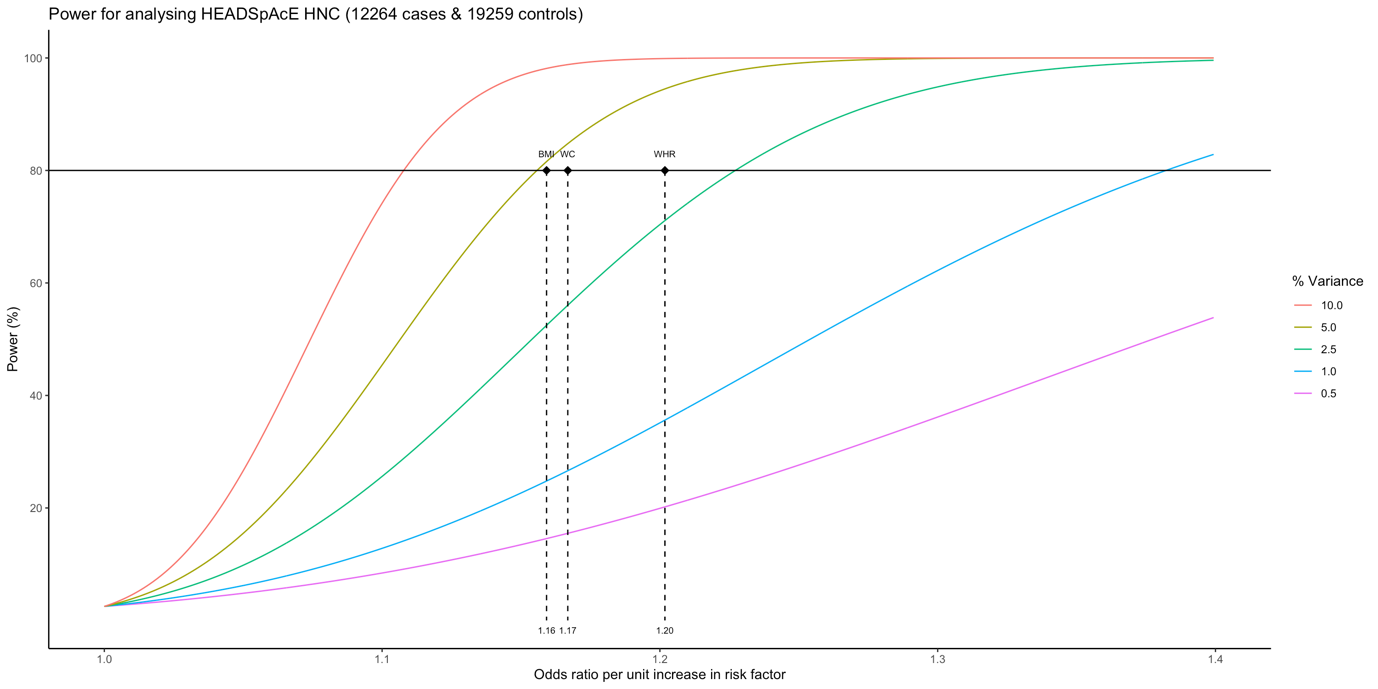

Supplementary Figure 1. Power calculations for HEADSpAcE head and neck cancer risk. Minimum odds ratios for body mass index (BMI), waist circumference (WC) and waist-to-hip ratio (WHR) were estimated assuming an alpha value of 0.05, power of 80% and total variances (R^2^) of 4.8%, 3.1% and 4.4%, respectively.

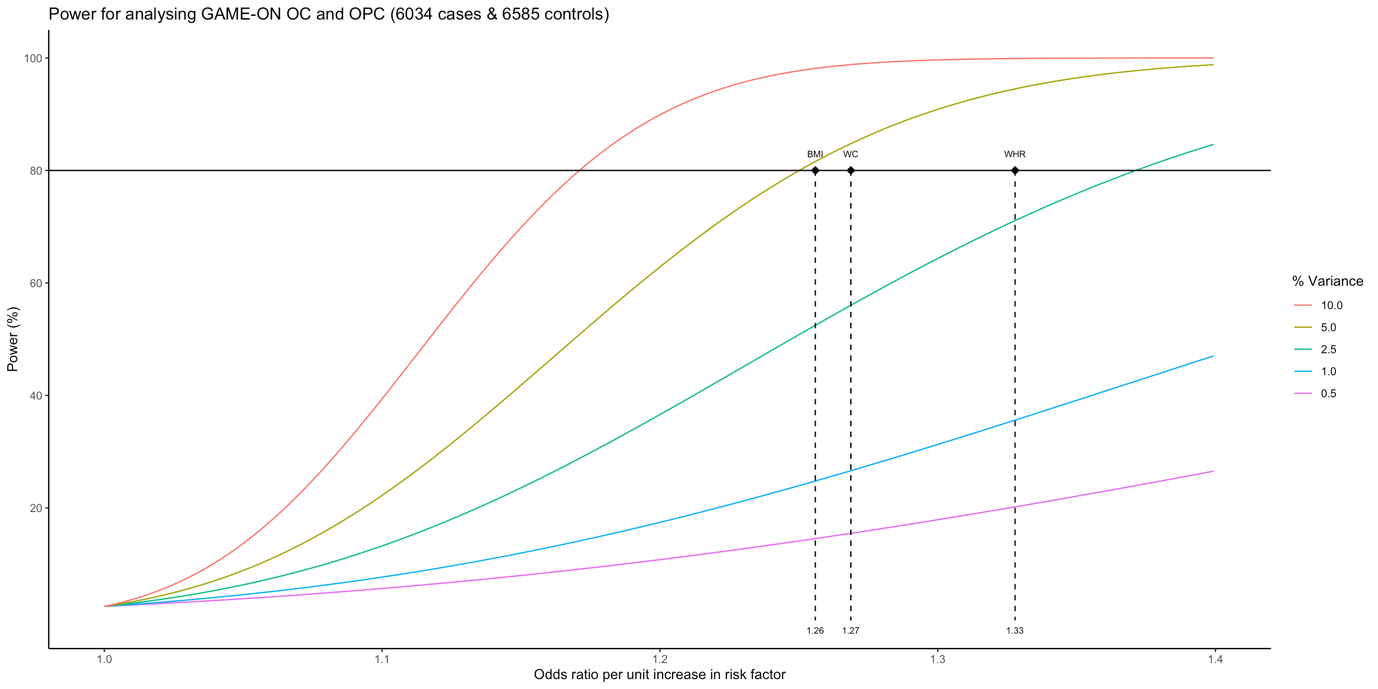

Supplementary Figure 2. Power calculation plot for GAME-ON oral cancer and oropharyngeal cancer risk. Minimum odds ratios for body mass index (BMI), waist circumference (WC) and waist-to-hip ratio (WHR) were estimated assuming an alpha value of 0.05, power of 80% and total variances (R^2^) of 4.8%, 3.1% and 4.4%, respectively.

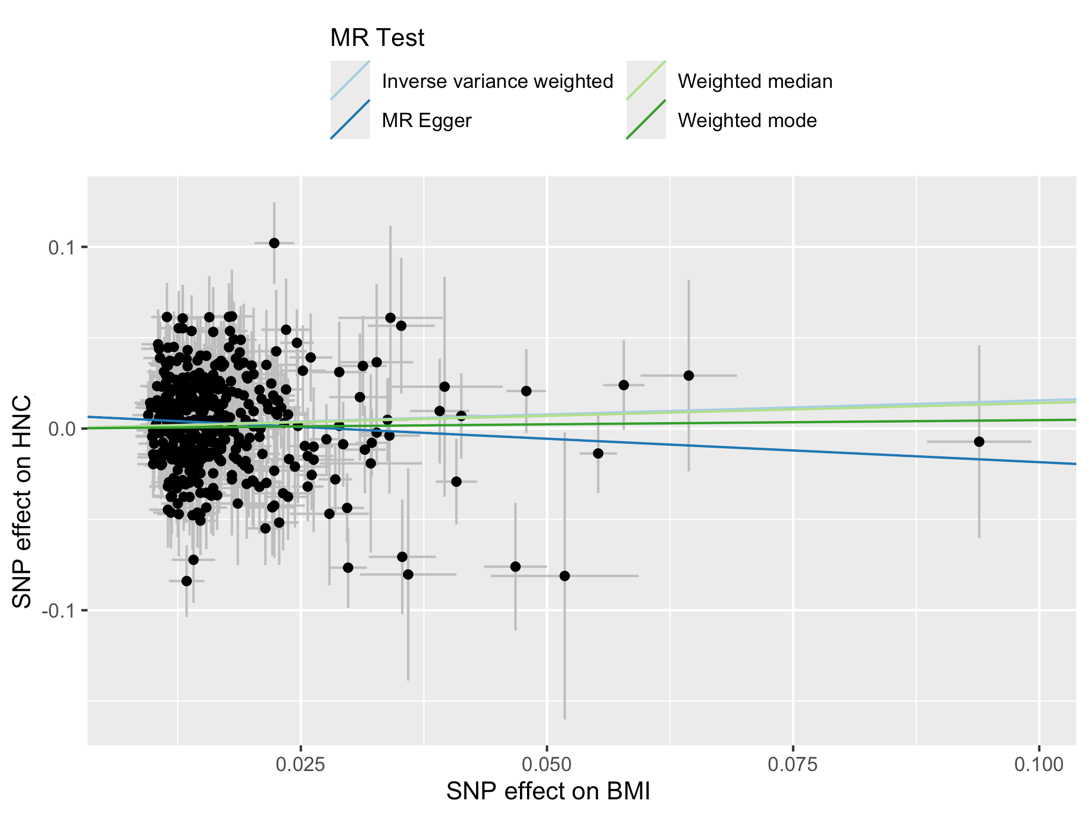

Supplementary Figure 3. Scatter plot for the genetically predicted effects of body mass index on the risk of head and neck cancer.

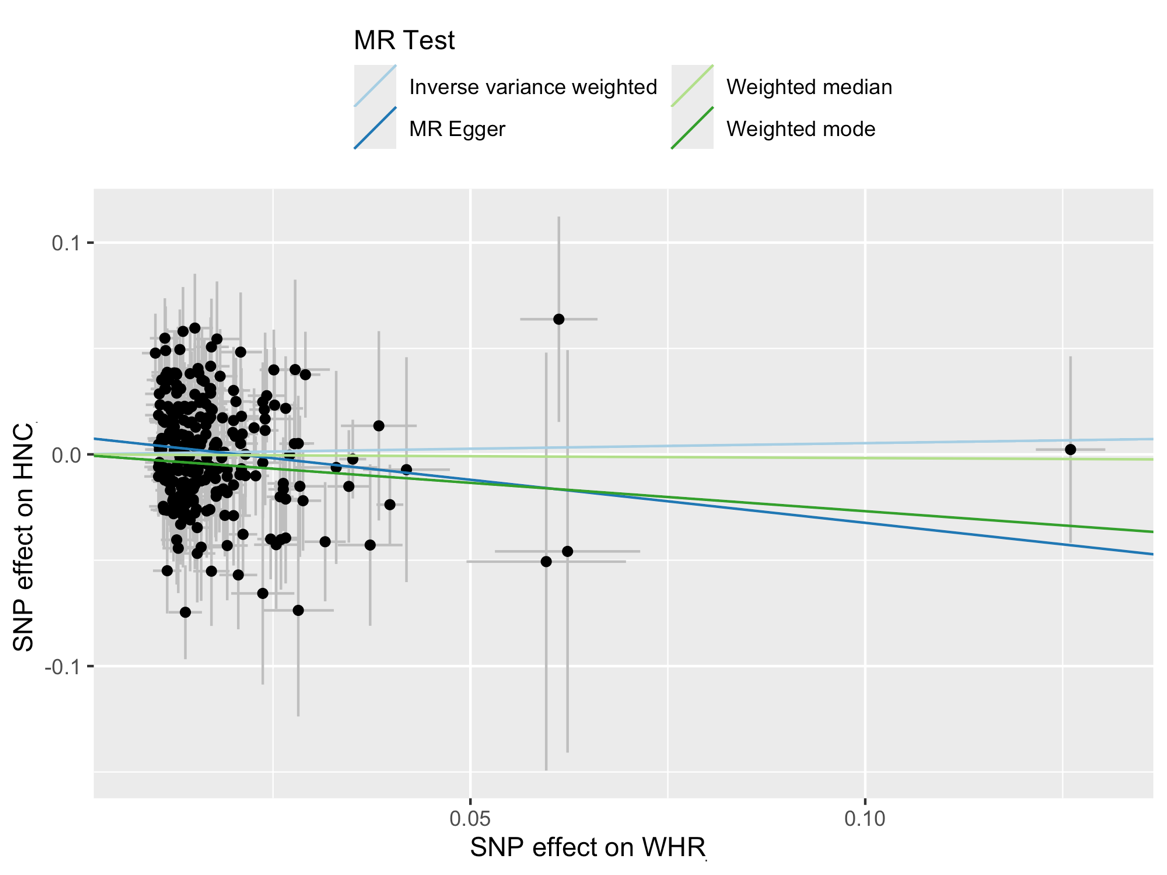

Supplementary Figure 4. Scatter plot for the genetically predicted effects of waist-to-hip ratio on the risk of head and neck cancer.

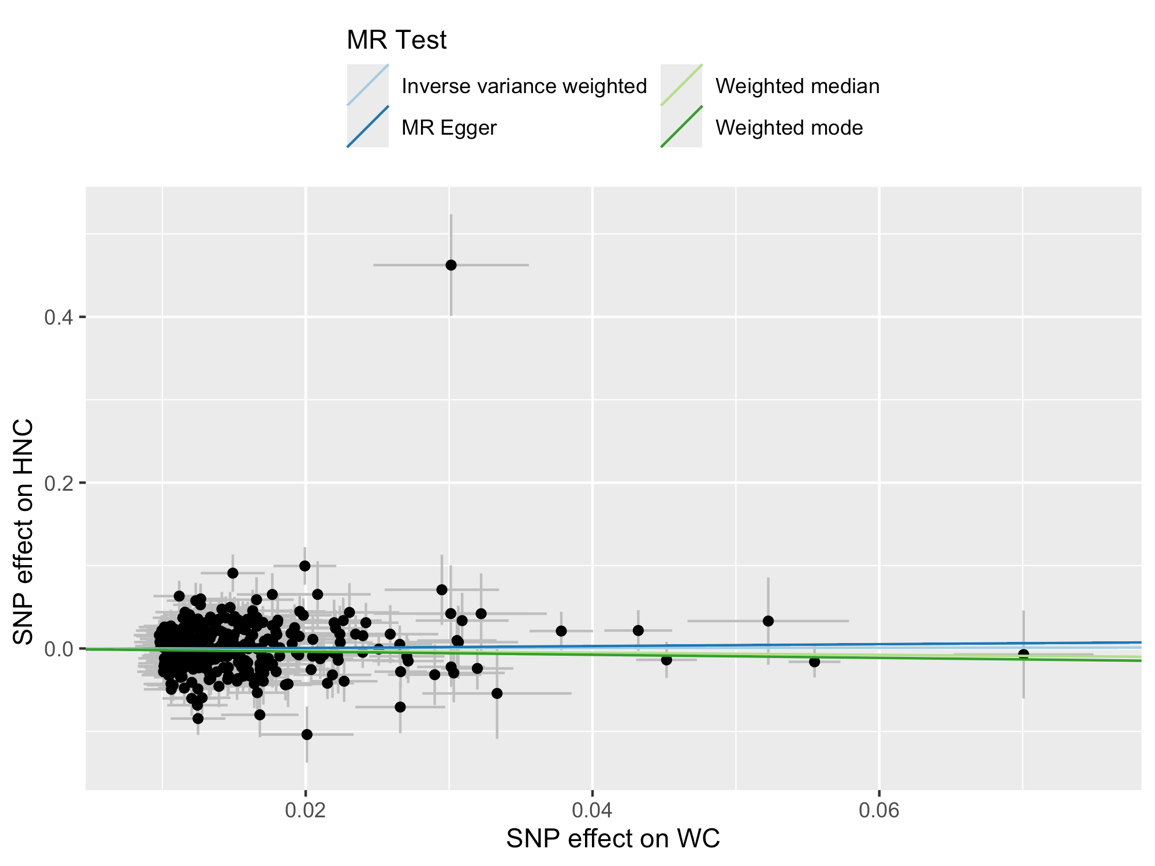

Supplementary Figure 5. Scatter plot for the genetically predicted effects of waist circumference on the risk of head and neck cancer.

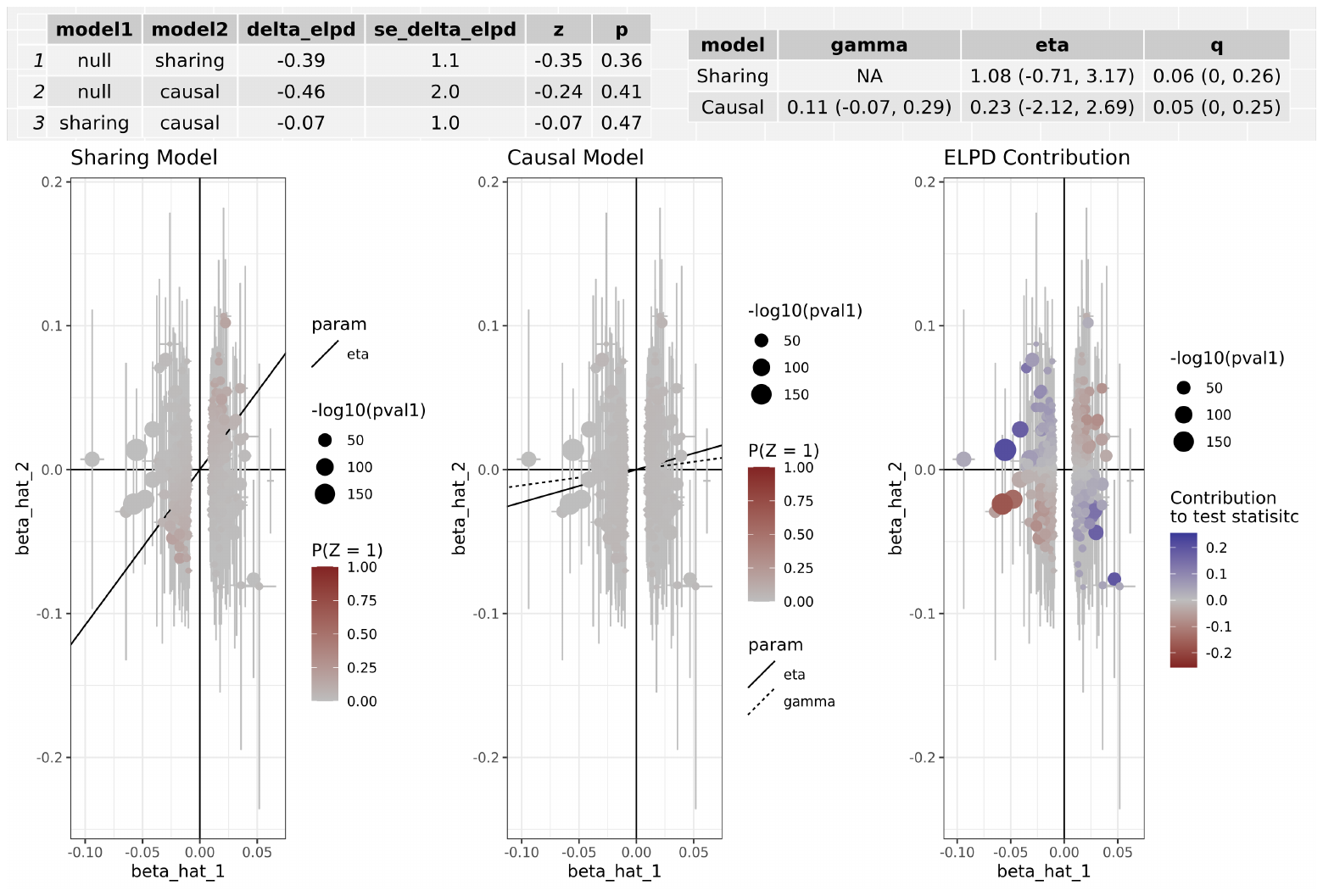

Supplementary Figure 6. CAUSE analysis for the genetically predicted effect of BMI on HNC risk. CAUSE estimates for HNC reported per 1-SD higher BMI in log odds ratio scale. The ELPD Contribution plot shows the relative contribution of each SNP to the CAUSE test statistic. Only SNPs with P < 5e-8 are shown. SNPs represented by larger circles reflect smaller p-values for the associations between genetic variants and BMI. SNPs that contribute more to the causal model are shown in warmer tones (i.e. red), while those that contribute more to the sharing model are shown in colder tones (i.e. blue). The delta_elpd is the statistic used to compare models. It is equal to elpd(model 1)- elpd(model 2). In the table on the left, negative delta_elpd’s suggest that model 2 is a better fit to the data than model 1 (i.e. that the sharing model is better than the null model in row 1, that the causal model is better than the null model in row 2, and that the causal model is better than the sharing model in row 3). The corresponding p-values test whether model 2 is a better fit than model 1. Here, row three suggests that the causal model is not a much better fit than the sharing model (the delta_elpd is negative but the p-value is 0.47, so there is no overwhelming evidence against the null hypothesis that the causal model is better than the sharing model). In the table on the right, eta represents the sharing factor effect (SNPs affect shared factor and shared factor simultaneously affects BMI and HNC) and gamma represents the causal factor effect (SNPs affect BMI and BMI affects HNC). Here, “0.11 (-0.07, 0.29)” represents the genetically predicted effect of BMI on HNC after adjusting for correlated and uncorrelated horizontal pleiotropy (results in log odds ratio scale). The intervals shown are credible intervals.

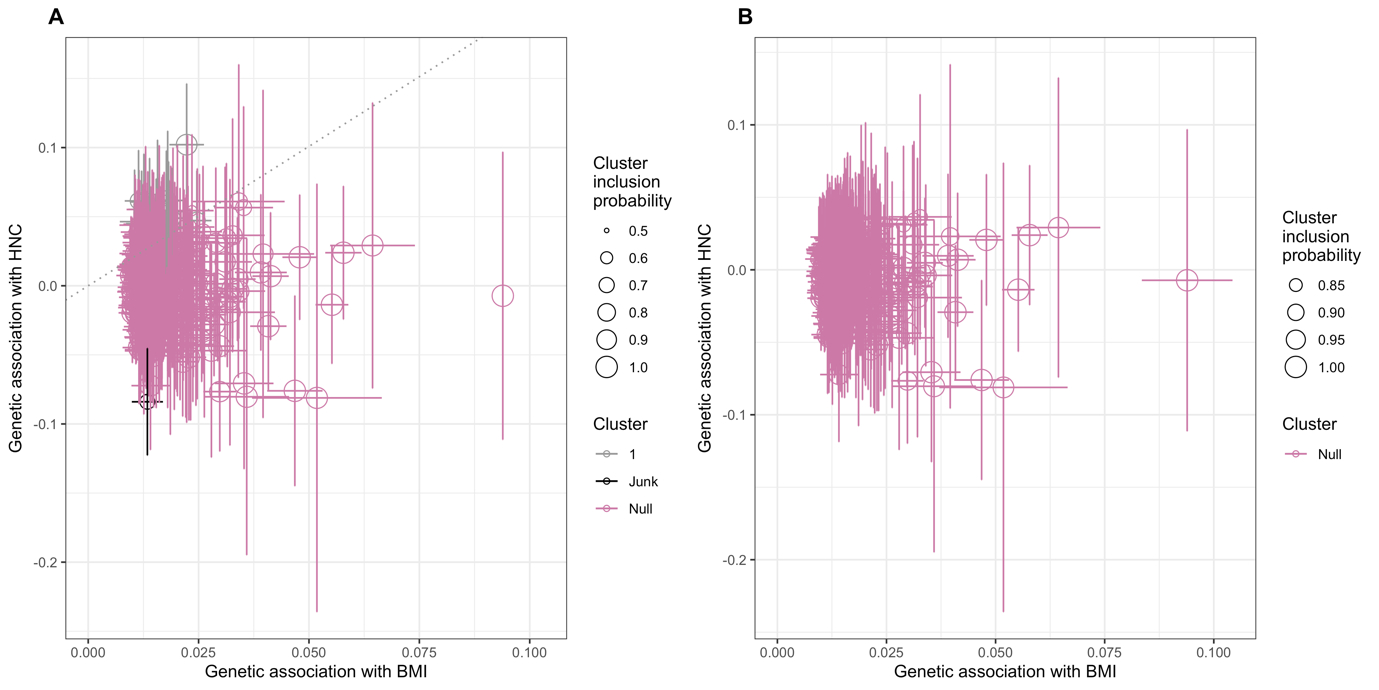

Supplementary Figure 7. Scatter plots depicting clusters of genetic associations with BMI and HNC, before (A) and after (B) conditional probability filtering.

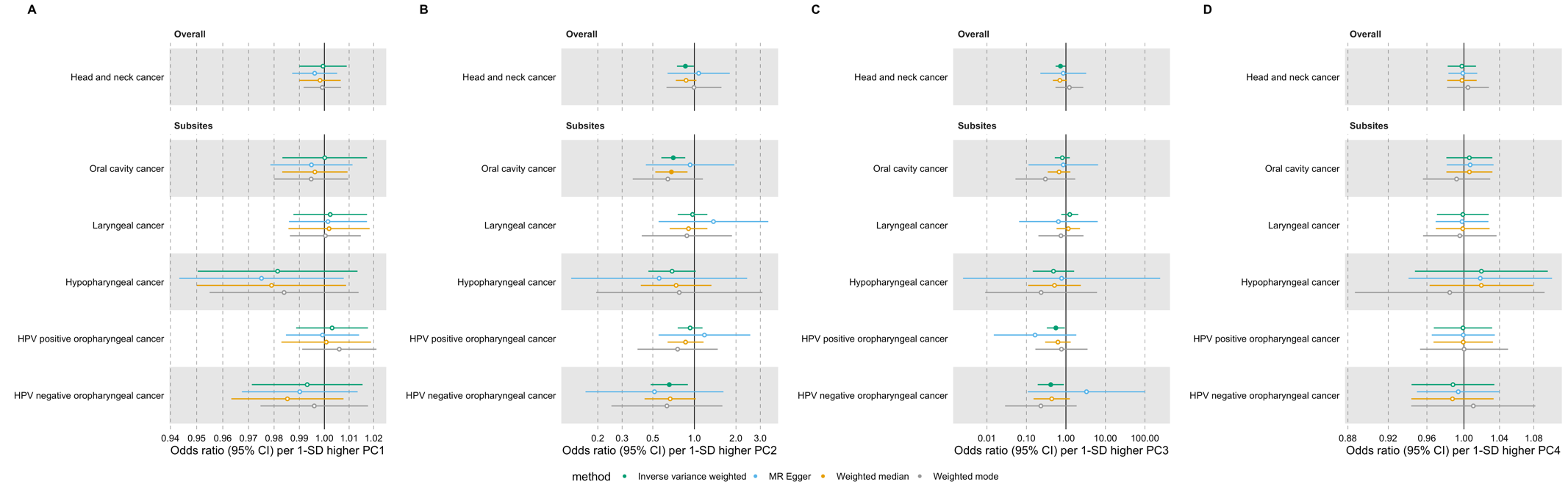

Supplementary Figure 8. Forest plots for the genetically predicted effects of four body shape principal components (PCs) on the risk of head and neck cancer and its subsites, where (A) PC1 is a measure of overall adiposity, (B) PC2 is a measure of tall and slim vs short and plump, (C) PC3 is a measure of tall with small hip vs short with big hip and (D) PC4 is a measure of high body mass index (BMI) and weight with small hip and waist vs low BMI and weight with big hip and waist.

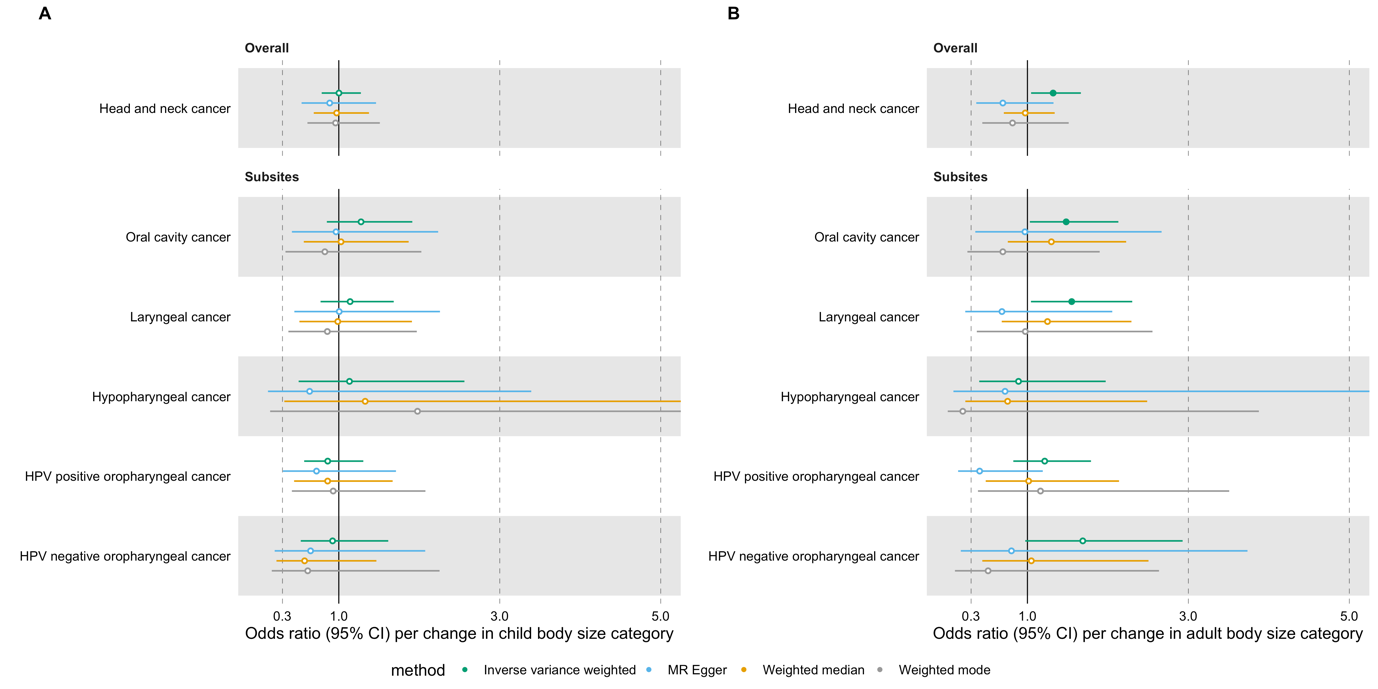

Supplementary Figure 9. Forest plots for the genetically predicted effects of (A) childhood and (B) adulthood body size on the risk of head and neck cancer and its subsites.

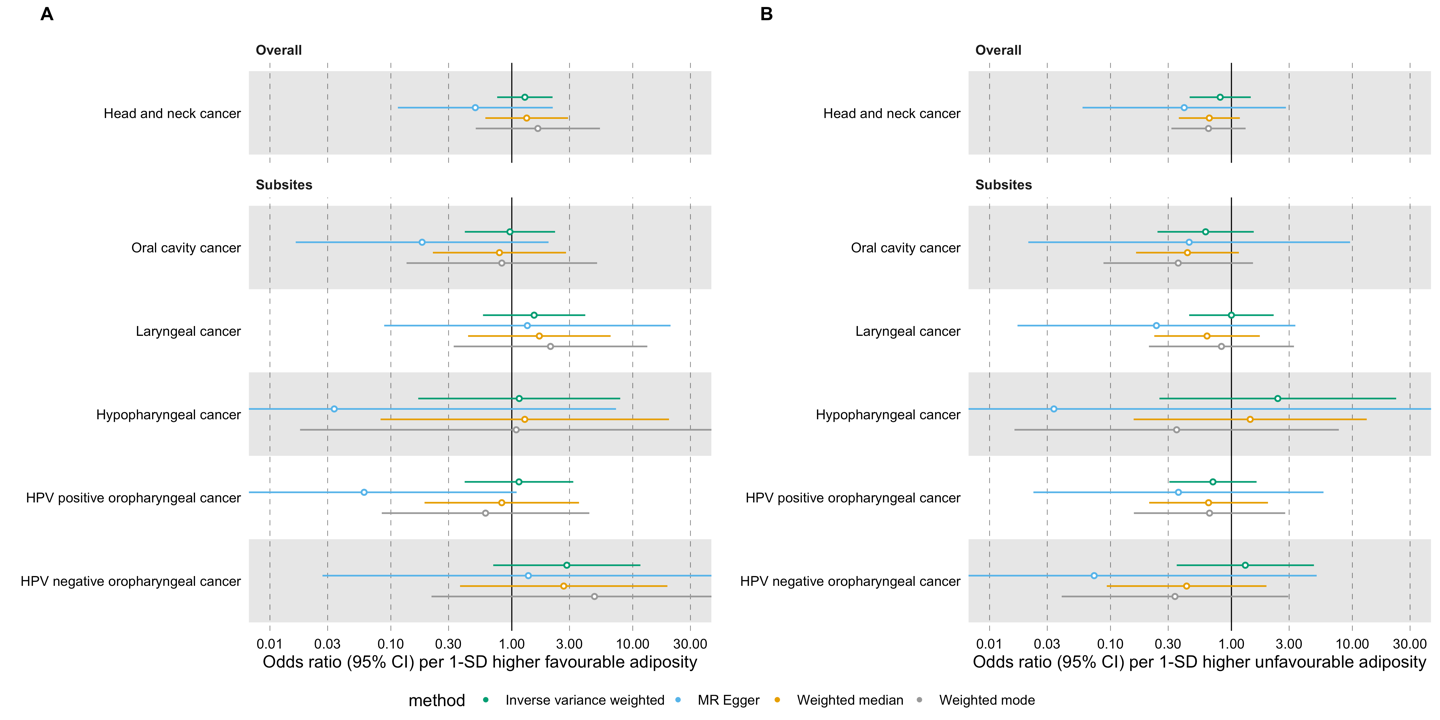

Supplementary Figure 10. Forest plots for the genetically predicted effects of (A) favourable and (B) unfavourable adiposity on the risk of HNC and its subsites.

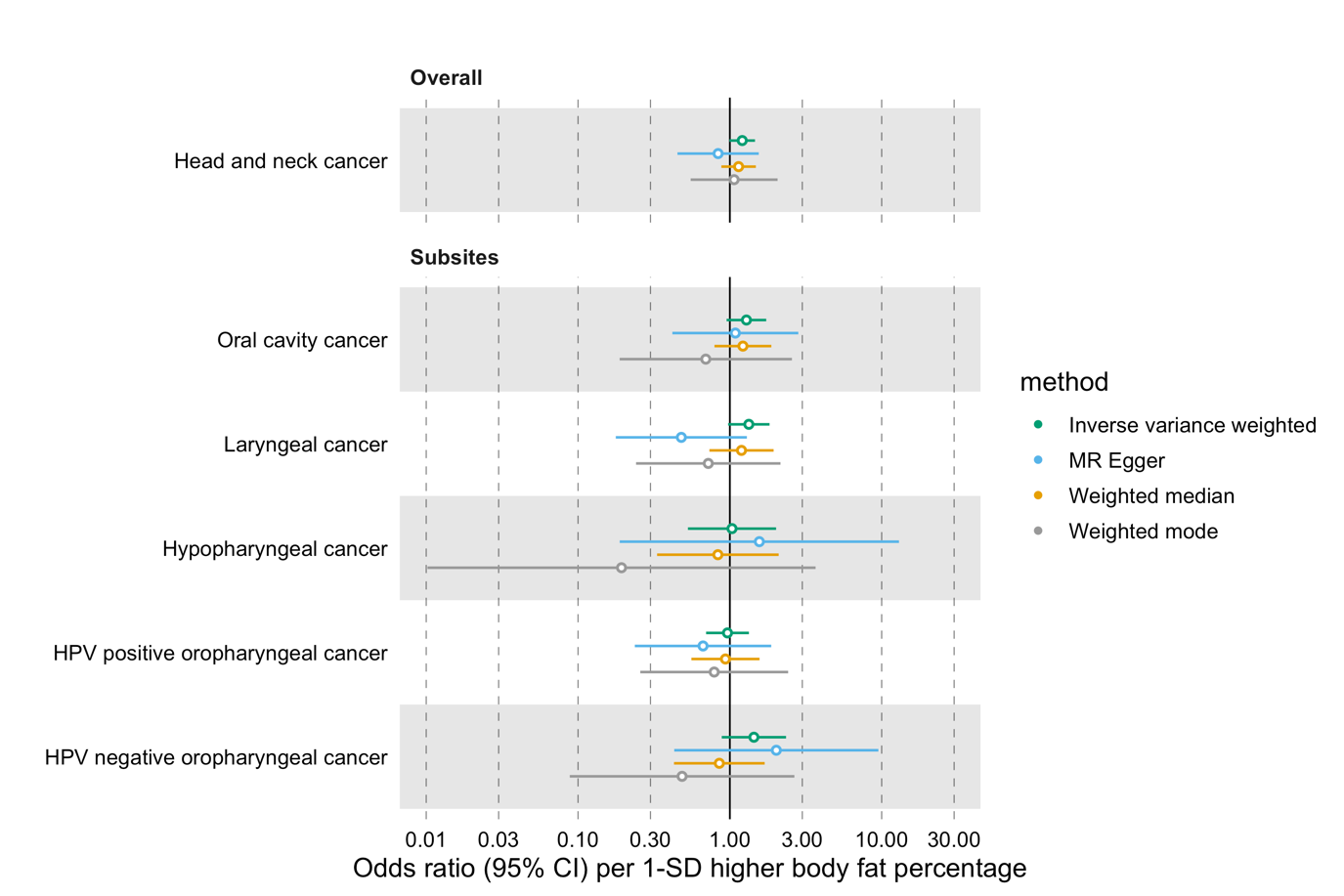

Supplementary Figure 11. Forest plots for the genetically predicted effect of body fat percentage on the risk of HNC and its subsites.

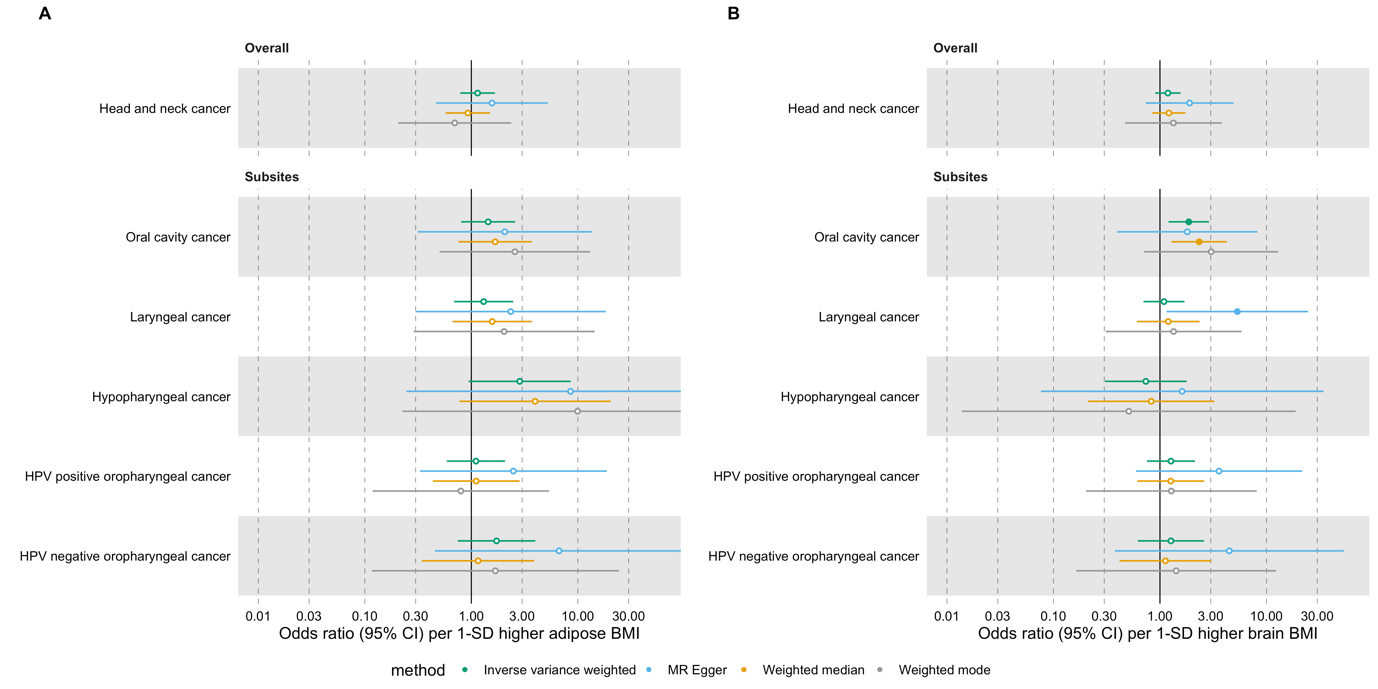

Supplementary Figure 12. Forest plots for genetically predicted effects of (A) adipose and (B) brain tissue-specific BMI on HNC and its subsites
